# Ankle and Toe Brachial Index Extraction from Clinical Reports For Peripheral Artery Disease Identification: Unlocking Clinical Data through Novel Methods

**DOI:** 10.1101/2021.05.08.21256421

**Authors:** Julia E. Friberg, Abdul H. Qazi, Brenden Boyle, Carrie Franciscus, Mary Vaughan-Sarrazin, Dax Westerman, Olga V. Patterson, Sharidan K. Parr, Michael E. Matheny, Shipra Arya, Kim G. Smolderen, Brian C. Lund, Glenn T. Gobbel, Saket Girotra

## Abstract

**Importance:** Despite its high prevalence and poor outcomes, research on peripheral artery disease (PAD) remains limited due to the poor accuracy of billing codes for identifying PAD in health systems.

**Objective:** Design a natural language processing (NLP) system that can extract ankle brachial index (ABI) and toe brachial index (TBI) values and evaluate the performance of extracted ABI/TBI values to identify patients with PAD in the Veterans Health Administration (VHA).

**Design, Setting, Participants:** From a corpus of 392,244 ABI test reports at 94 VHA facilities during 2015-2017, we selected a random sample of 800 documents for NLP development. Using machine learning, we designed the NLP system to extract ABI and TBI values and laterality (right or left).

Performance was optimized through sequential iterations of 10-fold cross validation and error analysis on 3 sets of 200 documents each, and tested on a final, independent set of 200 documents.

Performance of NLP-extracted ABI and TBI values to identify PAD in a random sample of Veterans undergoing ABI testing was compared to structured chart review.

**Exposure:** ABI ≤0.9, or TBI ≤0.7 in either right or left limb was used to define PAD at the patient-level

**Main Outcome:** Precision (or positive predictive value), recall (or sensitivity), F-1 measure (overall measure of accuracy, defined as harmonic mean of precision and recall)

**Results:** The NLP system had an overall precision of 0.85, recall of 0.93 and F1-measure of 0.89 to correctly identify ABI/TBI values and laterality. The F-1 measure was similar for both ABI and TBI (0.88 to 0.91). Recall was higher for ABI (0.95 to 0.97) while precision was higher for TBI (0.94 to 0.95). Among 261 patients with ABI testing (49% with PAD), the NLP system achieved a positive predictive value of 92.3%, sensitivity of 83.1% and specificity of 93.1% to identify PAD when compared to a structured chart review.

**Conclusion:** We have successfully developed and validated an NLP system to extract ABI and TBI values which can be used to accurately identify PAD within the VHA. Our findings have broad implications for PAD research and quality improvement efforts in large health systems.

## INTRODUCTION

Lower extremity peripheral artery disease (PAD) is a common and highly morbid condition,^1^ affecting an estimated 12.5 million people in the U.S. PAD is associated with high mortality, and is a leading cause of disability due to the associated risk of cardiovascular (CV) and limb events.^2-8^ The economic burden of PAD is enormous – the total annual cost of PAD due to hospital care alone was >$21 billion in 2004,^9-11^ and these costs are likely higher in 2021.

Contemporary research on PAD using electronic health record (EHR) data within large integrated health systems has been limited by the poor sensitivity (<40%) and specificity (60-70%) of billing codes for identifying PAD patients.^12, 13^ In contrast, the ankle-brachial index (ABI), a commonly used test for PAD diagnosis, has a high sensitivity (∼80%) and specificity (96%) for identifying PAD, which can be further enhanced with the addition of toe-brachial index (TBI).^14^ Moreover, ABI values represent an objective measure of leg ischemia, are associated with symptom severity and risk of CV events, and also influence treatment decisions (e.g., revascularization). ^15-19^ However, information on ABI and TBI values is commonly embedded in free text of the ABI test reports in the EHR and not directly available for research in most health systems. Manual extraction of ABI values can be time-consuming, limited by fatigue among human annotators and not practical in large health systems.

We developed and validated a natural language processing (NLP) system to extract ABI and TBI values from test reports obtained from 94 Veterans Affairs (VA) facilities. The NLP system uses machine learning-generated random forest models trained specifically to extract ABI and TBI values, assign laterality (right or left), and distinguish current from historical values from ABI test reports. We also evaluated the performance of an algorithm based on NLP-extracted ABI and TBI values to identify PAD in Veterans undergoing ABI testing.

## METHODS

### Setting and context

The study was conducted in the Veterans Health Administratin (VHA), which comprises 130 VA medical centers. The VA Corporate Data Warehouse (CDW) is a national repository that includes EHR data from all clinical encounters. Within the VHA, ABI tests are either performed in primary care, vascular surgery, or radiology depending on each site. Therefore, depending on the clinical department that performs the test, ABI test reports can be found in either Text Integration Utility (TIU) or Radiology domain in the CDW. We obtained data for all Veterans who had a TIU or a Radiology note entered in the CDW between 2015-2017. This work was approved by the Institutional Review Board and Research & Development Committee at both the Iowa City and Tennessee Valley VA.

### NLP System Design

#### Document Selection

We designed the NLP system to extract ABI and TBI values from ABI test reports rather than general clinical documents (e.g., clinic or consult notes) because ABI test reports represent the “source document” for these results in the EHR corresponding to the test performed on a given date. This approach permits the use of the date of the first abnormal ABI value as a surrogate for date of PAD onset, eliminates errors due to transcribing ABI data from test reports to clinical notes and avoids the ambiguity in distinguishing current ABI value from historical values which are commonly found in clinical notes. The overall approach is outlined in **Figure 1**. Details of the search strategy for identifying document titles used for reporting ABI test results from the Radiology and the TIU domain are included in the Appendix (**eMethods Section 1**.**1 & eTable 1**). A total of 224 document titles were identified from 94 out of 130 (72%) VA facilities which represented 392,244 documents on 222,033 unique patients during 2015-2017 in the VHA. We also found that some VA sites used a generic document title to report ABI and other vascular studies (e.g., ‘Vascular Studies’ note title for reporting all vascular studies including carotid, renal, mesenteric, along with ABI). We included documents associated with such generic titles in our final corpus for NLP training and testing to ensure that our tool could differentiate ABI test reports from other non-ABI vascular studies that use the same document title. Documents used for reporting ABI results could not be identified from 36 facilities primarily because they did not perform ABI testing (small facilities) or scanned ABI reports and stored them as images which are not available for research purposes)

**Figure 1.**
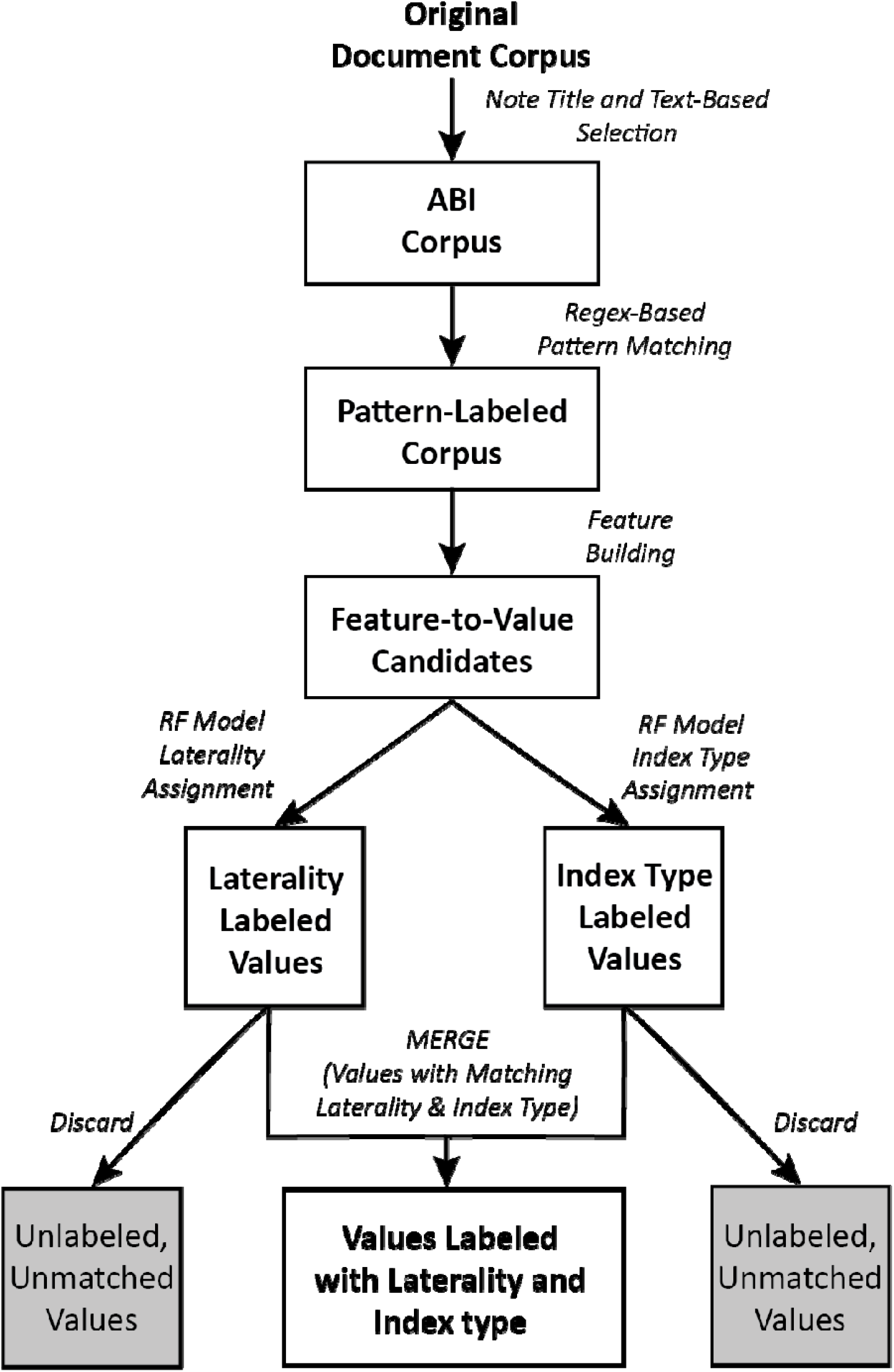
Flow diagram illustrating the steps used to extract ankle-brachial index values (ABIs) and toe-brachial index values (TBIs) from ABI testing reports generated within Department of Veterans Affairs medical facilities. The machine learning and NLP-based extraction system uses two, separately trained random forest (RF) models to assign index type (ABI, TBI, or neither) and laterality (left, right, or neither) to all values ranging from 0.0 to 2.0 in the document. Only values assigned to a specific index type and laterality are retained.

#### Annotation

From the total corpus of 392,244 ABI test reports during 2015-2017, we selected a random sample of 800 documents. Randomization was stratified to ensure that at least 5 documents were selected from each facility, with the remainder weighted by each facility’s volume of ABI test reports. The 800 reports were then separated into 4 sets of 200 documents each. A total of 600 documents were used for iterative NLP development and training and the remaining 200 documents were used for final testing.

Annotation was performed by two human annotators (JEF and AHQ). The goal of the annotation was to identify and categorize index type (ABI or TBI), index values, and laterality (right or left). Some ABI reports also included segmental blood pressure ratios, i.e., the ratio of blood pressure at the thigh or calf with arm blood pressure. Reviewers annotated these as ‘non-index’ values and the NLP system was deisgned to differentiate them from index (ABI or TBI) values using features of the surrounding text (e.g. “thigh index”). Reviewers also annotated ABI or TBI values in the reports that represent values from a prior study as “historical values”. All annotations were performed using eHOST software. ^20^ Additional details regarding annotations are included in the Appendix (**eMethods Section 1**.**2**)

#### NLP Development & Evaluation

The NLP tool uses regular expressions to label phrases commonly associated with the concept of ABI or TBI, their values, and laterality and uses a machine learning approach (random forest) for identification of the above concepts. A major challenge that we encountered was that ABI and TBI values were often reported in a semi-structured format (e.g., table). A rule-based approach, which relies on the proximity of the concept of ABI and TBI to its corresponding value and laterality is limited in a tabular format where the ABI values are often distant from the concept of ABI (usually a table row or a column header). To overcome this challenge, we designed our NLP system to search for ABI or TBI values that may be in vertical (column) or horizontal (row) alignment with the concept of ABI and TBI, and laterality, rather than just in close proximity of their respective values. Details of the NLP system design are included in the Appendix (**eMethods Section 1**.**3 and eTable 2**).

We adapted source code provided by the Weka Workbench to develop, train, and evaluate random forest models.^21^ We have made our code publicly available on Github.^22^ Two distinct models were trained, one for assigning index type and another for assigning laterality to candidate index values. Only values that were positively classified with respect to both index type (ABI or TBI) and laterality (left or right) were categorized as predicted index values (**Figure 2**). Random forest model hyperparameters were manually adjusted for each model using a grid search to maximize performance, which was quantified by comparing index values identified by the tool to those in the reference standard.

**Figure 2.**
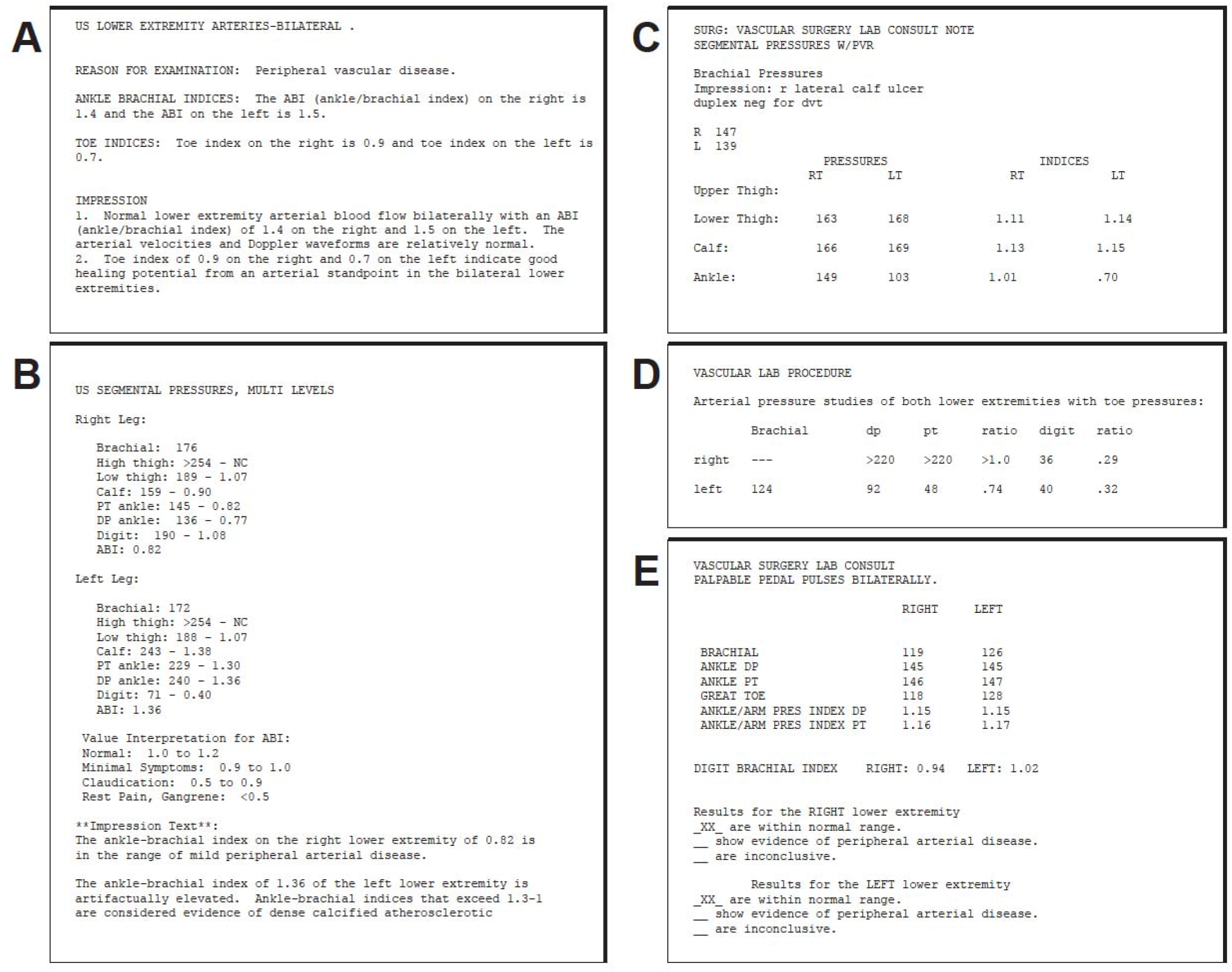
Five samples of ABI test reports illustrating the range of formats used for documenting ankle-brachial index values (ABIs) and toe-brachial index values (TBIs). There was substantial variation report format. In some (**A**), values were embedded in standard sentences, but ABIs and TBIs were commonly displayed using semi-structured, tabular formats that employed spaces and newline characters to define rows and columns (**B-E**). Various forms of row labels, column headings, and subheadings were used to indicate index type (ABI or TBI), laterality (left or right), and multiple other forms of information, such as pressures (**B-E**) and pressure ratios from the calf, thigh or other anatomical regions (**B&C**). All samples shown are from electronic health records of the Department of Veterans Affairs and were used in the present study for testing of the ABI and TBI extraction tool.

We employed an iterative process for tool development. After generating initial text patterns for feature extraction, we trained a random forest model on the first set of 200 documents and evaluated the trained model on a second set of 200 documents. During each iteration, we measured performance of the NLP tool against manual annotations performed by our study team. At each step, we conducted a detailed error analysis of false negative and false positive errors when compared to the reference standard, which was used to adjust the labeling patterns and features. We used cross-validation to test the modified system on training and evaluation set combined. This iterative process of evaluation followed by error analysis and system modification was continued until the change in F1-measure between two sequential cross validations run on independent testing sets was less than 1%. Once iterative development was complete, we evaluated tool performance on the final set of 200 documents.

Each ABI test report often had multiple mentions of ABI and TBI values (e.g., body of the report and conclusion). We designed the NLP system to extract all mentions of right ABI, left ABI, right TBI and left TBI in the annotated reference standard. Extracted values were considered as true positive if they exactly matched with the annotated set with regards to the index value, index type (ABI or TBI), *<u>and</u>* laterality (right or left). An extracted value was considered false positive if it did not match the annotated set on either the index value (e.g., 0.6 vs. 0.65), index type (ABI mislabelled as TBI) *<u>or</u>* laterality (e.g., right mislabelled as left). A false negative error was present when the NLP system completely missed an annotated value. We calculated precision (equivalent to positive predictive value), recall (equivalent to sensitivity) and F1-measure (harmonic mean of precision and recall) separately for mentions of right ABI, left ABI, right TBI and left TBI. We decided a priori that a precision, recall and F1-measure of >0.8 would be acceptable.^23^

#### Validation of the NLP system at a patient-level

After completing development and validation of the NLP system at a document level, we also assessed the performance of the NLP-extracted ABI and TBI values to identify patients with PAD when compared to structured chart review as the gold standard (criterion validity). For this step, we identified a random sample of 360 patients with an ABI test report (20 patients each from the 18 Veterans Integrated Service Networks [VISN]). Patients were included if the ABI test report was dated between January 1, 2016-June 30, 2017. As noted previously, many VA facilities use a common document title to report ABI and non-ABI vascular studies. Since non-ABI vascular studies were incorported in NLP training, we also evaluated the ability of the NLP system to differentiate patients who underwent non-ABI vascular studies. Details of the cohort inclusion criteria are listed in the Appendix (**eMethods Section 1**.**4**)

##### Chart Review

After initial training by the senior author (SG), structured chart review was performed by two physician trainees (AHQ and BB). Medical records dating back 5 years prior to the first ABI, and 6 months after the last ABI were selected for review. Data was extracted on demographics (age, sex, race), symptoms of PAD, diagnostic testing including other vascular imaging studies, and invasive treatment in a pre-designed chart abstraction form. Patients were classified as having lower extremity PAD if they met any one of the following criteria a) evidence of PAD on a confirmatory imaging study (e.g., angiography) b) physician documentation of PAD diagnosis in the EHR c) initiation or planned treatment of PAD (e.g., revascularization), d) prior history of PAD (e.g., revascularization), e) active leg symptoms in a patient with abnormal ABI but did not return for follow up. Patients were classified as no PAD if a) there was no evidence of PAD on confirmatory imaging study b) physician documentation of no PAD in the EHR, or c) no confirmatory testing performed in a patient with normal ABI (0.91 – 1.40). As noted above, ABI and TBI values alone were not used to determine PAD diagnosis during chart review except when patients with normal ABI who did not return for follow up visit. To ensure reliability, a random sample of 25 charts that were reviewed by the two trainees were independently reviewed by the senior author (SG). There was 100% agreement on the diagnosis of PAD variable between the chart reviewer and the senior author.

##### ABI Criteria for PAD

Based on the NLP-extracted ABI and TBI values, each patient was classified as having PAD if the ABI value was ≤0.9, or TBI value was <0.7 in any limb. The algorithm for PAD classification using the NLP extracted ABI and TBI values is displayed in **eFigure 1**.

## RESULTS

There was substantial variation in ABI test reports across VA facilities as illustrated in **Figure 2A-2E**. Although some ABI test reports contained largely complete, grammatical sentences (**2A**), most reports presented ABI and TBI values using a semi-structured format with a mixture of column headings, subheadings, and row labels that associate values to index type and laterality (**2B-E**). Additionally, some reports contained multiple values that appeared similar to but did not represent ABI or TBI such as 1) ratio of thigh to arm blood pressure (**2B & 2C**), 2) historical ABI or TBI values, or 3) a reference range for interpretation of index value ranges.

During the initial testing and cross validation of the NLP system on the first set of 200 documents, the overall precision was 0.72, recall was 0.55, and F-1 measure was 0.63. Iterative development, which involved error analysis and modification of regular expressions for text pattern recognition resulted in gradual performance improvement as the development set was expanded to encompass a total of 600 documents. Once development was complete and no further improvements were gained by further changes to the system, precision, recall, and F1-measure combined across index type and laterality was 0.92, 0.94, and 0.93, respectively **(Table 1)**. Addition of each set of location-dependent features resulted in incremental improvement in overall precision, recall, and F1-measure, with the highest performance achieved with the inclusion of all feature sets (**eTable 3)**.

**Table 1.** Cross-Validation-Based Performance of the ABI Extraction Tool on the Development Set.

| **Index Type and Laterality** | **TP** | **FP** | **FN** | **Precision** | **Recall** | **F1** |
| --- | --- | --- | --- | --- | --- | --- |
| *ABI Right* | 467 | 42 | 20 | 0.92 | 0.96 | 0.94 |
| *ABI Left* | 449 | 37 | 29 | 0.92 | 0.94 | 0.93 |
| *TBI Right* | 142 | 15 | 10 | 0.90 | 0.93 | 0.92 |
| *TBI Left* | 128 | 10 | 13 | 0.93 | 0.91 | 0.92 |
| *Total* | 1186 | 104 | 72 | 0.92 | 0.94 | 0.93 |

*TP* – true positive mentions of index values; *FP* – false positive mentions; *FN* – false negative mentions

The final performance of the NLP assessed is shown in **Table 2**. Overall precision was 0.85, recall was 0.93, and the F1-measure was 0.89. In some cases, the tool identified ABI values missed by the annotators, and, after correction for these oversights, overall precision, recall, and F1-measure were slightly higher at 0.86, 0.94, and 0.90, respectively. The F1-measure of the NLP system exceeded the inter-annotator agreement of the human reviewers (Appendix, **eTable 4**). The F1-measure was similar regardless of index type (ABI or TBI) or laterality (right or left). and ranged from 0.88 to 0.91, but precision and recall varied by the index type. Recall was higher for ABI than TBI values, while precision was higher for TBI compared to ABI. Details regarding the false positive and false negative errors on the NLP system are included in the Appendix (**eResults**).

**Table 2.** Performance of Random Forest-based ABI Extraction Tool on the Final Testing Set.

| **Index Type and Laterality** | **TP** | **FP** | **FN** | **Precision** | **Recall** | **F1** |
| --- | --- | --- | --- | --- | --- | --- |
| *ABI Right* | 111 | 22 | 4 | 0.83 | 0.97 | 0.90 |
| *ABI Left* | 108 | 24 | 6 | 0.82 | 0.95 | 0.88 |
| *TBI Right* | 35 | 2 | 5 | 0.95 | 0.88 | 0.91 |
| *TBI Left* | 32 | 2 | 6 | 0.94 | 0.84 | 0.89 |
| *Total* | 286 | 50 | 21 | 0.85 | 0.93 | 0.89 |

*TP* – true positive mentions of index values; *FP* – false positive mentions; *FN* – false negative mentions

### Patient-level validation

Of the 360 patients randomly selected for the standardized chart review cohort, 99 (27.5%) patients had a non-ABI vascular study. The NLP system correctly identified all of the 99 test reports as non-valid ABI, and therefore they were excluded from the chart review. The characteristics of the remaining 261 patients are displayed in **Table 3**. The mean age was 69.6 years and 18.9% were Black, 5% were Hispanic. There was a high prevalence of co-morbidities in this cohort including hypertension, diabetes, chronic kidney disease, and heart failure.

**Table 3.** Characteristics of Chart Review Cohort.

| Variable | N=261 |
| --- | --- |
| Mean Age, years (SD) | 69.6 (9.9) |
| Male Sex | 256 (98.1%) |
| Race* |  |
| White | 200 (76.6%) |
| Black | 49 (18.9%) |
| Other | 6 (2.3%) |
| Hispanic Ethnicity* | 13 (5.0%) |
| Symptoms |  |
| Claudication | 68 (26.1%) |
| Atypical leg pain | 60 (23.0%) |
| Rest pain or ulcer | 46 (17.6%) |
| None documented | 87 (33.3%) |
| Known history of PAD | 53 (20.3%) |
| Co-morbidities |  |
| Hypertension | 237 (90.8%) |
| Diabetes | 149 (57.1%) |
| Chronic Kidney disease | 83 (31.8%) |
| COPD | 117 (44.8%) |
| Heart failure | 77 (29.5%) |
| Prior history of MI | 29 (11.1%) |
| Cerebrovascular disease | 75 (28.7%) |
| Cancer | 68 (26.1%) |
| ABI test results, Mean (SD) |  |
| Right ABI | 0.97 (0.28) |
| Left ABI | 1.00 (0.25) |
| Right TBI | 0.66 (0.28) |
| Left TBI | 0.66 (0.24) |

Numbers in the table represent N(%) or Mean (SD).
**Abbreviations:** COPD: Chronic Obstructive Pulmonary Disease; MI: Myocardial Infarction; SD: standard deviation
\*Race was unknown for 6 patients, ethnicity was unknown for 4 patients

A total of 130 (49.8%) patients were categorized as PAD on chart review. The NLP system classified 117 (44.8%) patients as PAD and 121 (46.3%) patients without PAD. For 23 (8.8%) patients, the NLP system was unable to extract any ABI or TBI values (9 patients with PAD and 14 patients without PAD on chart review). In the primary analysis, we included these patients and classified them as no PAD. The positive predictive of the NLP system for identifying PAD was 92.3%, sensitivity was 83.1% and specificity was 93.1% (Table 4). In the first post-hoc sensitivity analysis, we assumed that the 23 patients who couldn’t be classified using the NLP system were completely misclassified, i.e., patients with PAD on chart review classified as No PAD using NLP and vice versa (i.e., worst case scenario). In this analysis, the positive predictive value was 82.4%, sensitivity was 83.1% and specificity was 82.4%. Excluding the 23 patients above (second post-hoc sensitivity analysis) yielded a positive predictive value of 92.3%, sensitivity of 89.3%, and specificity of 92.3%.

**Table 4.**
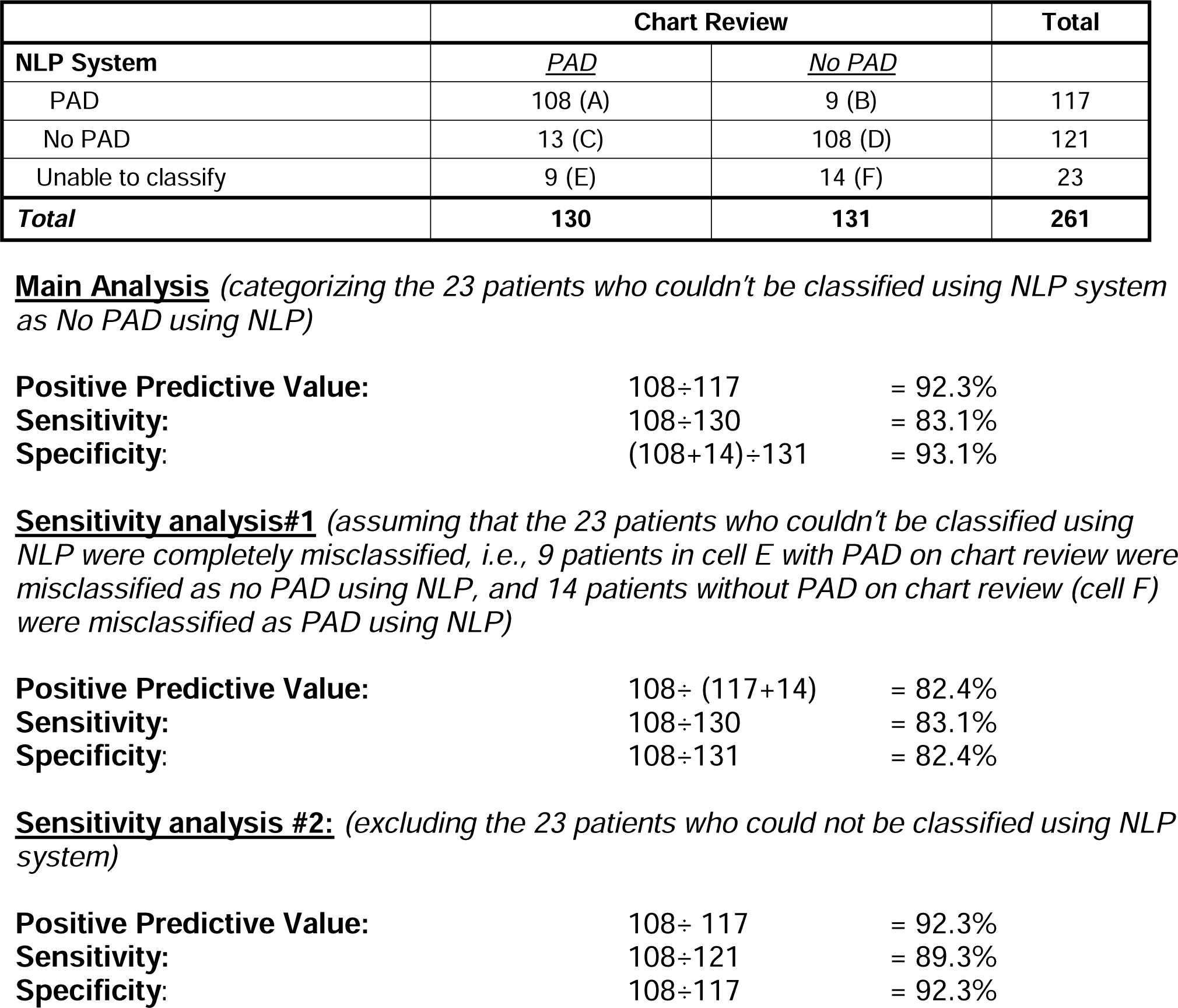
Performance of the NLP system in identifying patients with PAD compared to structured chart review.

| NLP System | Chart Review |  | Total |
| --- | --- | --- | --- |
|  | <u>PAD</u> | <u>No PAD</u> |  |
| PAD | 108 (A) | 9 (B) | 117 |
| No PAD | 13 (C) | 108 (D) | 121 |
| Unable to classify | 9 (E) | 14 (F) | 23 |
| **Total** | **130** | **131** | **261** |

**Main Analysis** (categorizing the 23 patients who couldn't be classified using NLP system as No PAD using NLP)
|  |  |  |
| --- | --- | --- |
| **Positive Predictive Value:** | $108 \div 117$ | = 92.3% |
| **Sensitivity:** | $108 \div 130$ | = 83.1% |
| **Specificity:** | $(108+14) \div 131$ | = 93.1% |

|  |  |  |
| --- | --- | --- |
| **Positive Predictive Value:** | $108 \div (117+14)$ | = 82.4% |
| **Sensitivity:** | $108 \div 130$ | = 83.1% |
| **Specificity:** | $108 \div 131$ | = 82.4% |

**Sensitivity analysis #2:** (excluding the 23 patients who could not be classified using NLP system)
|  |  |  |
| --- | --- | --- |
| **Positive Predictive Value:** | $108 \div 117$ | = 92.3% |
| **Sensitivity:** | $108 \div 121$ | = 89.3% |
| **Specificity:** | $108 \div 117$ | = 92.3% |

## DISCUSSION

Using ABI test reports from 94 VA facilities that varied substantially in terms of document structure and complexity, we have successfully developed and validated an NLP system that can extract ABI and TBI values with a high degree of accuracy, with an overall precision of 0.85, recall of 0.93 and an F1-measure of 0.89. Furthermore, an algorithm based on NLP-extracted ABI and TBI values had a positive predictive value of 92.3% in identifying patients with PAD. The successful development of an NLP system for PAD detection in the VHA, the largest integrated health system in the U.S., provides an unprecedented opportunity to advance research and quality improvement efforts for PAD – a common and highly morbid condition that accounts for a substantial clinical and economic burden. Several of our findings are important and merit further discussion.

Traditional approaches that rely on billing codes for identifying PAD patients have been limited by poor sensitivity (<40%) and specificity (60-70%), which have hampered PAD research and quality improvement efforts.^12, 13^ Accordingly, there has been a growing interest in developing NLP systems for PAD identification and many different approaches have been described. ^24-27^ In a recent study, Weissler et al developed an NLP system using the EHR data at a single academic medical center.^24^ The NLP system determined the probability of PAD based on ABI values, imaging studies and prior history of revascularization or non-traumatic amputation and was superior to a prior model that used billing codes with an overall positive predictive value of 74%, specificity of 62%, when sensitivity was set at 90%. Other NLP studies for PAD identification have been conducted in highly selected populations (e.g., patients undergoing amputation^26^ or lower extremity angiograms^25^) and the utility of such models for broad clinical and investigational purposes remains unclear.

Our NLP system differs from prior applications which have primarily used a manual, rule-based approach that relies on the proximity of the concept of ABI/TBI with their corresponding values and laterality.^26^ During the initial review of ABI test reports, we found that many documents reported ABI and TBI values in a tabular format. Extracting ABI and TBI values from such data require consideration of the complex interaction between column headings, sub-headings, and row labels which would make a manual rule-based model challenging.^28^ To address this challenge, we designed our NLP system using a random forest-based, machine learning model, which is able to detect the relationship between the relative location of text patterns and their interaction with ABI and TBI values. A major innovation of our NLP sysem is that vertical and horizontal alignment of text patterns rather than proximity alone which ensured a high degree of accuracy in identify ABI and TBI values.

In addition to the accuracy of the NLP system in extracting individual ABI and TBI values, an algorithm based on the above values achieved high performance in identifying patients with PAD with an overall positive predictive value of 92.3% sensitivity of 83.1% and specificity of 93.1%. Even under the assumption that the 23 (8.8%) patients in whom the NLP system could not extract any ABI values were completely misclassified, the performance of our NLP system exceeds that of billing codes which have a sensitivity <40% and specificity <70%. Our findings are consistent with prior population-based studies that have reported a high sensitivity (80%) and specificity (96%) of abnormal ABI (<0.9) for identifying PAD.^14^ Despite their accuracy, ABI and TBI values are not routinely available as discrete data elements in most EHRs, including the VHA and are often embedded within clinical documents as either unstructured text or containing semi-structured elements such as tables. Manual extraction of ABI and TBI is not feasible given the large volume of ABI tests (>50,000) performed each year in the VHA. Our NLP system will allow for obtaining information on ABI and TBI values efficiently, and the latter can be used for identifying patients with PAD with a high degree of accuracy.

Accurate automatic extraction of ABI and TBI values will permit identification of large cohort of PAD patients within the VA’s integrated health system. Although the tool achieved a high positive predictive value, its performance may be further enhanced in the future with the incorporation of findings from imaging studies. Since the NLP tool was developed and validated using VHA data, it can be readily scaled to the national VHA with high fidelity that will promote the use of big data analytics for PAD research. In addition, the NLP system can be intergrated into evaluating PAD care delivery, identifying opportunities for improving care quality and setting the stage for interventions including clinical trials to address current and emerging research questions. Thus, our findings have important implications for advancing PAD research and quality improvement efforts in the VHA.

The findings of our study should be interpreted in the context of the following limitations. First, even though we included documents from a large number of VA facilities, ABI test reports from ∼30% of VA facilities could not be obtained. Many of these facilities scan the ABI test reports and store them as images making them unavailable in the VHA CDW for research purposes. Alternative strategies such as developing algorithms to identify PAD using clinic notes at these sites maybe developed. Second, our study was conducted using VHA data and therefore the accuracy of our tool on documents outside the VA remains untested. However, given the large variation in document structure and complexity included in the study, we believe that retraining the model on non-VA documents might be sufficient for optimizing the NLP system for non-VA sites. Third, among patients with a valid ABI, the NLP system was unable to extract ABI and TBI values (and unable to classify PAD) in 8.8% patients (23 out of 261). Even under the assumption that these patients were completely misclassified, the performance of the NLP system exceeded that of PAD billing codes. Additional refinements to the NLP system may further reduce the proportion of patients in whom ABI and TBI values cannot be extracted. Fourth, our NLP system was designed using random forest models to automate the identification of important feature interactions. It is possible that additional performance gains may be achieved using more advanced models (e.g., deep learning) that have the advantage of partially automating the generation of features and feature interaction.^28-31^ Finally, given that the VHA guidelines do not recommend screening ABI in asymptomatic patients, patients with undiagnosed or asymptomatic PAD would not be included as they are unlikely to undergo ABI testing.

In conlusion, we have successfully developed and validated an NLP system that can extract ABI and TBI values and identify patients with PAD within the VA’s large integrated health system with a high degree of accuracy. Our work has important implications for advancing research and quality improvement efforts in the VHA and elsewhere.

## Data Availability

Due to the nature of this research, participants of this study did not agree for their data to be shared publicly, so supporting data is not available

