## Supplementary Appendix for "Ankle and Toe Brachial Index Extraction from Clinical Reports For Peripheral Artery Disease Identification: Unlocking Clinical Data through Novel Methods"

Center for Access and Delivery Research and Evaluation, Iowa City Veterans Affairs Medical Center (JEF, CF, MVS, BCL, SG) and Department of Medicine, University of Iowa Carver College of Medicine (MVS, SG), Iowa City, IA; Massachusetts General Hospital, Boston, MA (AHQ); Division of Cardioscular Medicine, University of Minnesota, Minneapolis, MN (BB); Salt Lake City Veterans Affairs Medical Center, Salt Lake City, UT (OVP); Tennessee Valley Healthcare System (MEM, GTG) and Vanderbilt University Medical Center (DW, SKP, MEM, GTG), Nashville, TN; Palo Alto Veterans Affairs Medical Center and Stanford University, Palo Alto, CA (SA); Department of Medicine and Psychiatry, Yale University School of Medicine, New Haven, CT (KGS)

*Julia E. Friberg and Abdul H. Qazi contributed equally to this manuscript

^$^ Glenn T. Gobbel and Saket Girotra contributed equally to this manuscript

**Co-corresponding Authors:**

**Saket Girotra MD, SM**

200 Hawkins Drive, Suite 4427 JCP
Division of Cardiovascular Diseases, University of Iowa

Iowa City, IA 52242

**Glenn T. Gobbel, DVM, PhD, MS**

4^th^ Floor, GRECC, Tennessee Valley Health System

1310 24th Ave S, Nashville, TN 37212


**eMethods**

**1.1 Identifying ABI Test Report Documents Within the VA Corporate Data Warehouse**

The VA Corporate Data Warehouse (CDW) includes clinical documents from many VA clinical and administrative systems and includes multiple domains. Documents that contain ABI test results can be found in either Text Integration Utility (TIU) or Radiology domain, depending on the clinical department that conducts the ABI test at a particular site.

In the Radiology domain, all procedural test results including ABI test reports are always associated with a Current Procedure Terminology (CPT) code. We used the linkage of the ABI CPT does listed in **eTable 1** to identify all ABI test reports in the Radiology domain. However, this strategy could not be used for the TIU domain because CPT codes are not consistently associated with an ABI test report in that domain. Moreover, each VA facility can define custom document titles that can include procedure names (e.g., [site name] ABI test report) within the title. Accordingly, to identify ABI test reports in the TIU domain, we performed a search on all documents in the TIU domain that contained a term corresponding to the ABI concept in either the document title or in the text of the document. From the identified document titles, we retrieved a a sample of corresponding documents to determine if they represented an ABI test report. A final list of all document titles that correspond to ABI test reports was curated. Using the above strategy, we identified a total of 224 document titles that correspond to an ABI test report from 94 VA facilities.

1.2 **Annotation of ABI Test Reports**

Two reviewers (AHQ and JEF) performed all annotations for the project. Initial reviewer training was conducted on a set of documents that were separate from the corpus of 800 documents selected for the study. After training, each of the two reviewers annotated distinct sets of 300 documents (600 total). All annotations were reviewed by the study senior authors (SG and GG). In regular team meetings, disagreements in annotations were reviewed and adjudicated by joint consensus, and the annotation guidelines were updated as needed. For the final set of 200 documents that was used for evaluation of the NLP system and measuring inter-annotator agreement (IAA), annotations were completed by both reviewers with final adjudication by the senior authors.

**1.3 Design of the NLP system**

The development of the NLP system was conducted in three sequential processes which included 1) text processing, 2) feature generation and 3) random forest-based machine learning Text processing first divided the document into sections based on commonly identified patterns, such as particular headers followed by newline characters. The sections were then split into sentences and tokens using open source, machine-learning trained models^21^ combined with our own custom rule set previously optimized for VA clinical notes. Finally, the system assigned all tokens to a sentence, a horizontal line of text, and one or more vertical columns of text. We defined the vertical columns of a particular document as the minimal set of lines, each one character in length, extending from the top and through the bottom line of text that would intersect all tokens in the document. For the purposes of this study, we considered all tokens intersected by the same vertical line to be “aligned.”

The feature generation module employed regular expressions to detect any numeric values between 0.00 and 2.00, and these were referred to as candidate indices. Then the context around the candidate indexes was encoded using a set of lexical features identified using regular expressions to label matched sequences with one of 13 categories and 37 subcategories (**eTable2)**. For example, the phrase “Ankle-brachial index” would be labeled as being within the “Index Value Type” category and “ABI” type subcategory. All feature definitions also included positional information in a sentence and text line, distance in characters and tokens from the index candidate, and alignment with other features. Alignment was included to account for the semi-structured elements in the reports and it captured not only horizontal but relative vertical positioning of features in text.

The machine learning module utilized the features associated with each annotated index value to train the random forest model, which was then used for classifying candidate index values with respect to index type, index laterality (left or right), and temporality (current or historical). Random forests consist of multiple decision trees generated through random subsampling of the data.^22^ During training, the machine learning process automatically integrates the multiple feature interactions, like those which may occur in text, into the random forest model.

**1.4 Patient-level Valiation of the NLP system**

Inclusion and Exclusion Criteria: Between January 1, 2016 to June 30, 2017, we identified all patients who had at least one ABI test. Patients were included only if their ABI test reports were not already included among the 800 ABI test reports previously identified for NLP development and training. We required that included patients should not have an ABI in the 12-month prior to enrich our cohort with patients undergoing ABI testing for new symptoms. Finally, we also restricted our cohort to patients age ≥ 40 years of age since PAD is rare in younger patients and patients with at least 2 outpatient, or 1 outpatient and 1 inpatient encounter to focus our study on regular VHA users. From this cohort, a random sample of 360 patients (20 patients from each VISN) were selected.

**eResults**

Inspection of overall performance showed a higher prevalence of false-positive (FP) labels than false-negative labels (50 vs. 21, **Table 3**). Within the 200-document test set, there were 501 “candidate” values with a form similar to ABI and TBI values and 307 of these were true, current ABI or TBI values with an assigned laterality. The remaining 194 values were numerical values between (0.00 to 2.00) that did not represent true ABI or TBI values (e.g., historical ABI values or ratio of thigh to arm blood pressure). Out of the 50 total FPs, 39 (78%) came from the remaining 194 candidate values. Erroneous labeling of historical values accounted for 34% of the FPs, and 44% were attributable to erroneous labeling of non-ABI/non-TBI values (e.g., brachial-calf pressure ratio), numbers indicating lists (e.g., ‘1.’, ‘2.’), or values within non-ABI reports. Incorrect assignment of index type (e.g., substitution of ‘ABI’ for ‘TBI’) accounted for 16% of the FPs, and 6% of the FPs were actually ABI and TBI values missed by the annotators, which were mentioned above.

**eFigure 1** Algorithm based on NLP-extracted ABI or TBI values to identify PAD

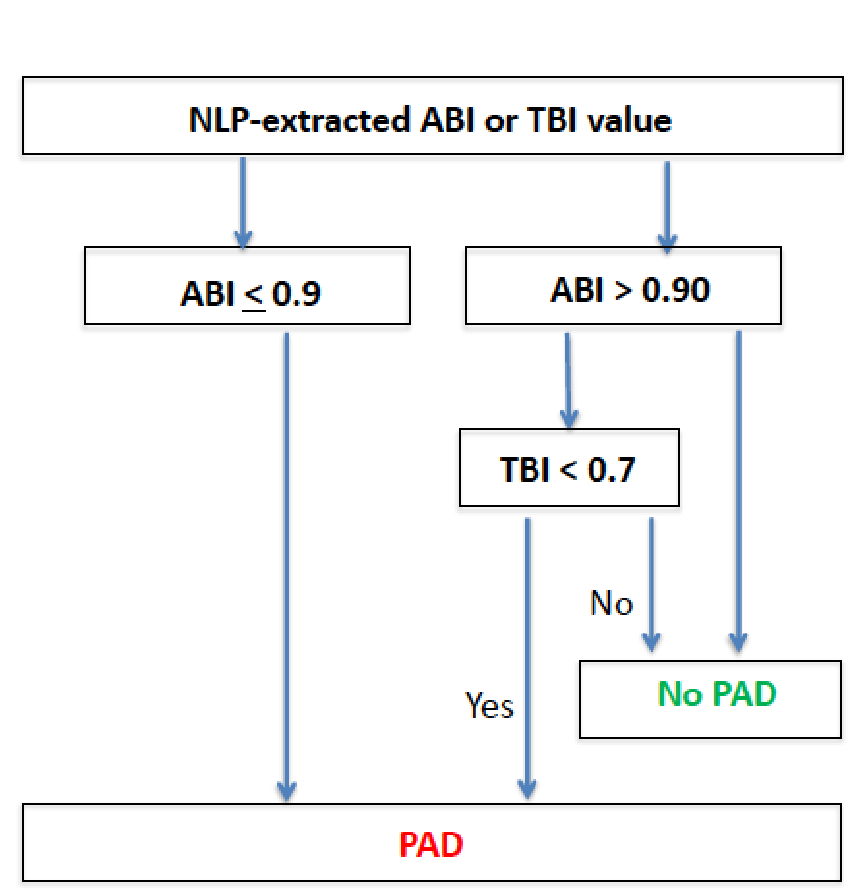

**eTABLES**

**eTable 1 List of CPT codes for ABI testing**

| **CPT Code** | **Description** |
| --- | --- |
| 93922 | Limited bilateral noninvasive physiologic studies of upper or lower extremity arteries, (eg, for lower extremity: ankle/brachial indices at distal posterior tibial and anterior tibial/dorsalis pedis arteries plus bidirectional, doppler waveform recording and analysis at 1-2 levels, or ankle/brachial indices at distal posterior tibial and anterior tibial/dorsalis pedis arteries plus volume plethysmography at 1-2 levels, or ankle/brachial indices at distal posterior tibial and anterior tibial/dorsalis pedis arteries with transcutaneous oxygen tension measurements at 1-2 levels) |
| 93923 | Complete bilateral noninvasive physiologic studies of upper or lower extremity level study with provocative functional maneuvers (eg, measurements with postural provocative tests, or measurements with reactive hyperemia) arteries, 3 or more levels (eg, for lower extremity: ankle/brachial indices at distal posterior tibial and anterior tibial/dorsalis pedis arteries plus segmental blood pressure measurements with bidirectional doppler waveform recording and analysis, at 3 or more levels, or ankle/brachial indices at distal posterior tibial and anterior tibial/dorsalis pedis arteries plus segmental volume plethysmography at 3 or more levels, or ankle/brachial indices at distal posterior tibial and anterior tibial/dorsalis pedis arteries plus segmental transcutaneous oxygen tension measurements at 3 or more level(s), or single |
| 93924 | Noninvasive physiologic studies of lower extremity arteries, at rest and following treadmill stress testing, (ie, bidirectional doppler waveform or volume plethysmography recording and analysis at rest with ankle/brachial indices immediately after and at timed intervals following performance of a standardized protocol on a motorized treadmill plus recording of time of onset of claudication or other symptoms, maximal walking time, and time to recovery) complete bilateral study |
| 93925 | Duplex scan of lower extremity arteries or arterial bypass grafts; complete bilateral study |

**eTable 2** – Categories and subcategories of regular expressions used to identify text patterns and generate features for machine learning using random forest models.

| **Category** | **Subcategories** |
| --- | --- |
| anatomy | ankle |
|  | arm |
|  | other |
|  | thighOrCalf |
|  | toe |
| artery | ankle |
|  | arm |
|  | unspecified |
| exercise | exercise |
| impression | impression |
| interpretation | interpretationHeader |
| indexValue | indexValue |
| indexValueType | abi |
|  | tbi |
|  | unspecified |
| laterality | bilateral |
|  | left |
|  | right |
| negativeIndexInfo | flow |
|  | indexValueInterpretation |
|  | listIndex |
|  | miscellaneous |
|  | nonIndexArtery |
|  | otherIndexType |
|  | pressureValue |
|  | unit |
| noncompressible | noncompressible |
| pressure | pressureAnkle |
|  | pressureToe |
|  | pressureUnspecified |
| range | bounded |
|  | unbounded |
| temporal | beginLineDate |
|  | date |
|  | historical |
|  | nonhistorical |
|  | time |

**eTable 3** Impact of Location-Dependent Features on ABI Extraction Tool Performance

| **Cross Validation Set** | **Model Features Included** | | | **Counts** | | | **ABI Tool Performance** | | |
| --- | --- | --- | --- | --- | --- | --- | --- | --- | --- |
|  | **Line** | **Column** | **Document** | **TP** | **FP** | **FN** | **Precision** | **Recall** | **F1** |
| 1 | **-** | **-** | **-** | 914 | 344 | 370 | 0.73 | 0.71 | 0.72 |
| 2 | **+** | **-** | **-** | 1116 | 142 | 132 | 0.89 | 0.89 | 0.89 |
| 3 | **-** | **+** | **-** | 1050 | 208 | 247 | 0.85 | 0.81 | 0.82 |
| 4 | **-** | **-** | **+** | 1141 | 117 | 159 | 0.91 | 0.88 | 0.89 |
| 5 | **+** | **+** | **-** | 1158 | 100 | 115 | 0.92 | 0.91 | 0.92 |
| 6 | **+** | **-** | **+** | 1175 | 83 | 115 | 0.93 | 0.91 | 0.92 |
| 7 | **-** | **+** | **+** | 1153 | 105 | 147 | 0.92 | 0.89 | 0.90 |
| 8 | **+** | **+** | **+** | 1176 | 82 | 109 | 0.94 | 0.92 | 0.92 |

***TP*** – true positive mentions of index values; ***FP*** – false positive mentions; ***FN*** – false negative mentions

+ model feature included; **-** model feature excluded

**eTable 4.** Interrannotator agreement for the Testing set

| **Index Type and Laterality** | **Agreements** | **Disagreements** | **F1** |
| --- | --- | --- | --- |
| *ABI Right* | 72 | 33 | 0.814 |
| *ABI Left* | 68 | 33 | 0.805 |
| *TBI Right* | 32 | 11 | 0.853 |
| *TBI Left* | 28 | 11 | 0.836 |
| *Total* | 200 | 88 | 0.820 |

***TP*** – true positive mentions of index values; ***FP*** – false positive mentions; ***FN*** – false negative mentions
